## Supplementary material for "Analogous humoral antigen recognition between Monkeypox-infected and Smallpox-vaccinated individuals"

**Supplementary figures**

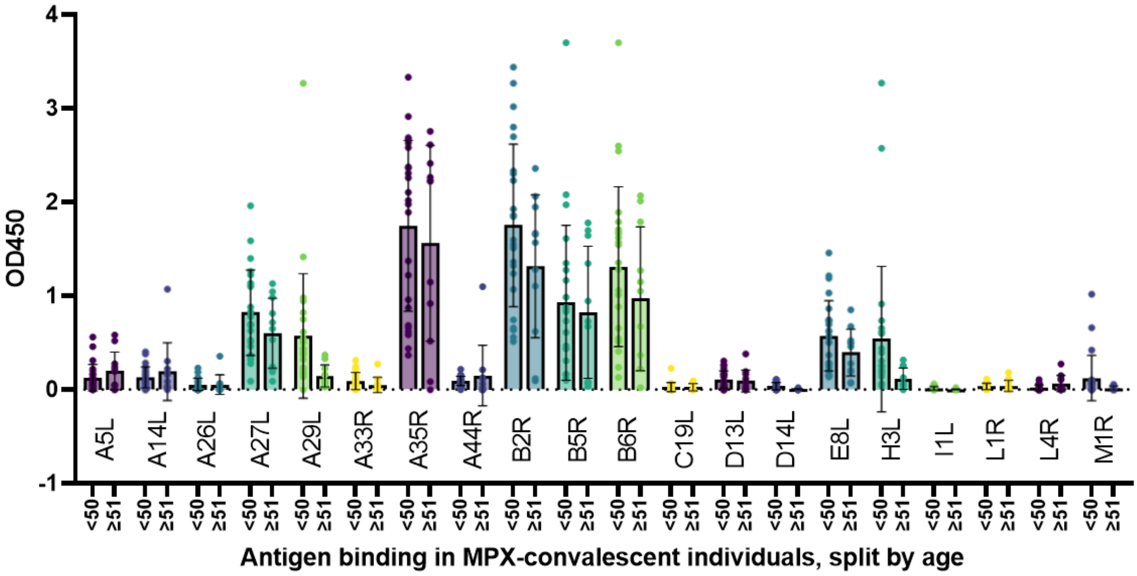


**Supplementary Figure 1**: Antigen binding in MPX individuals to different MPXV/VACV antigens, split according to age group (<50, >=51).

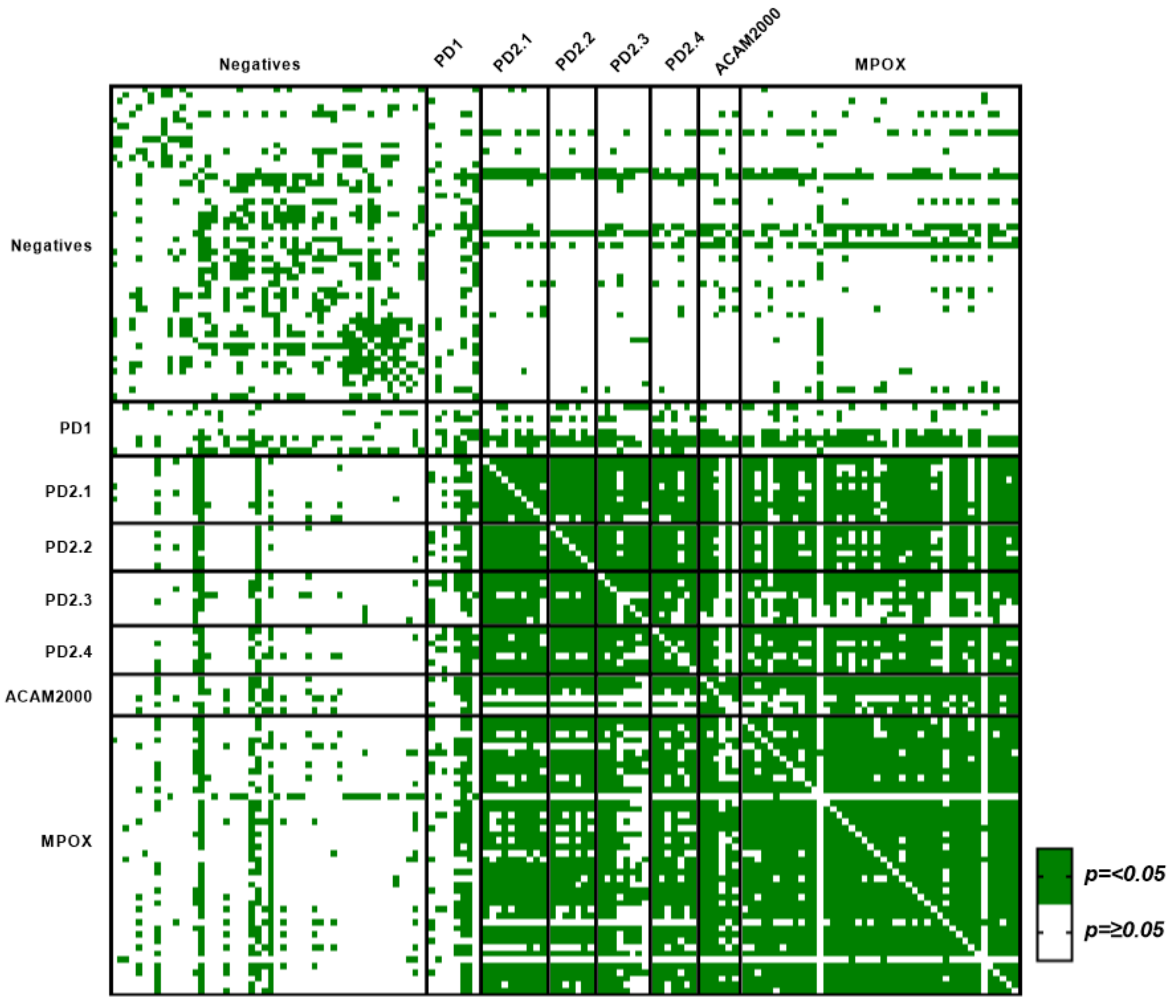


**Supplementary figure 2:** P-values of the Pearson correlation matrix. Green indicates a p value of <0.05.

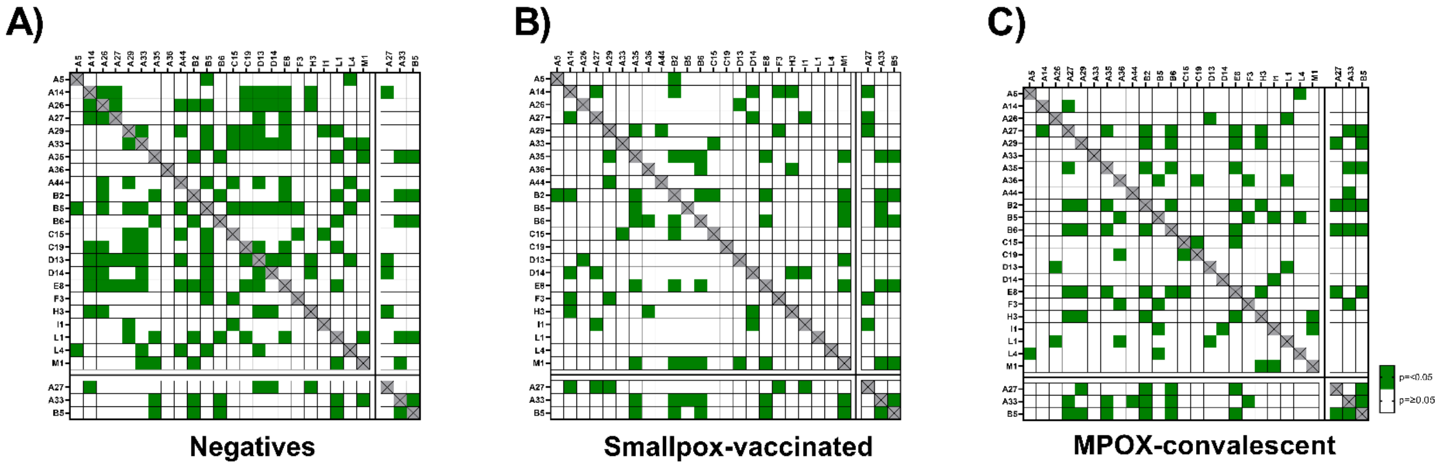


**Supplementary Figure 3:** P-values for MPXV/VACV antigens to one another in the different cohorts (negative, Smallpox-vaccinated and MPOX-convalescent). Gree

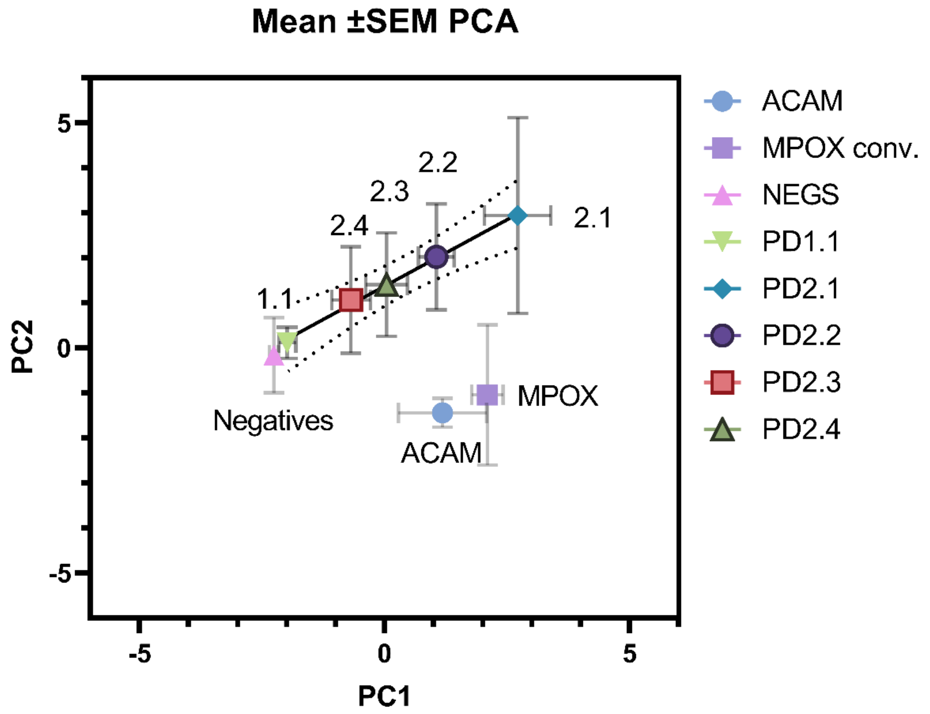


**Supplementary Figure 4:** Principal component analysis, with mean PCA +/- SEM for each group. Simple linear regression (solid black line) was applied for all post-Imvanex vaccination samples.

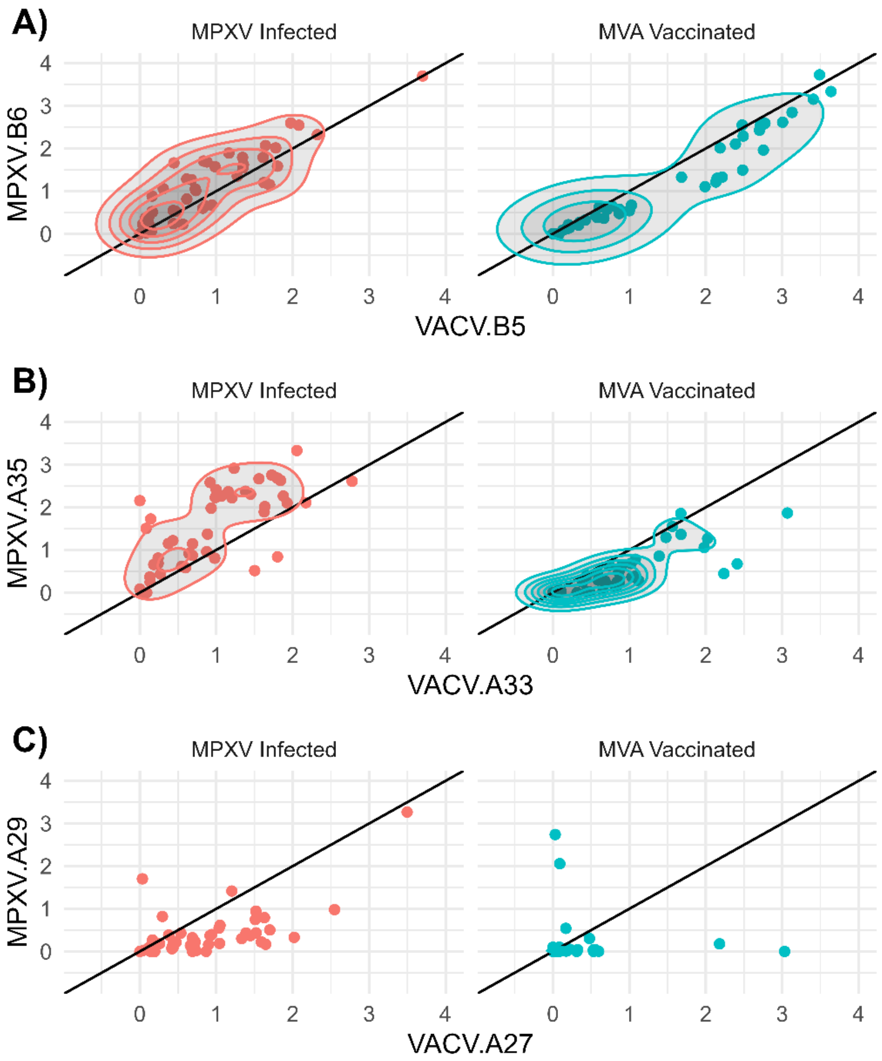


**Supplementary figure 5:** Correlation of binding between MPXV and VACV protein homologues in MPXV-infected or MVA-vaccinated individuals.

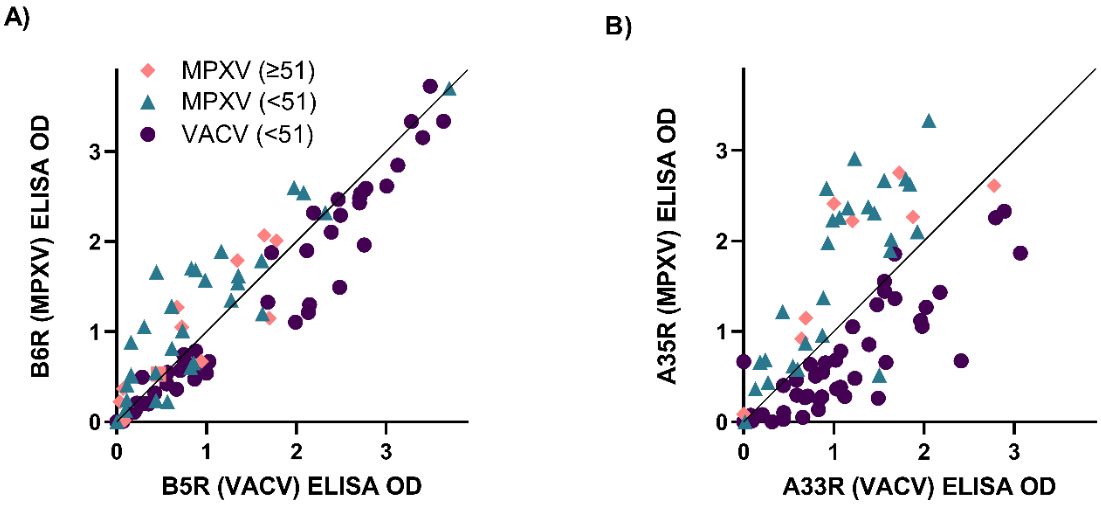


**Supplementary figure 6**: Antibody binding in MPXV-infected individuals aged <51 or >=51 to homologous VACV and MPXV antigens.

**Supplementary tables**

**Supplementary table 1:** Antigens used in this study, the viral source, company, expression host, expressed fragment and sequence provided by the manufacturer.

| MPXV/VACV Protein | Virus | Company | Expression Vector | Expressed Fragment | Sequence |
|---|---|---|---|---|---|
| **A5** | MPXV | Native Antigen Company | E. coli | Total protein | MDFFNKFSQG LAESSTPKSS IYYSEEKDPD TKKDEAIEIG LKSQESYYQR QLREQLARDN MMTASRQPTQ PLQPTIHITP QPVPTPTPAP ILLPSSTAPV LKPRQQTNTS SDMSNLFDWL STDTDAPAST LLPALTPSNT VQDIISKFNK DQKMTTPPST QPSQTLPTTT CTQQSDGSIS CTTPTVTPLQ PPIVATVCTP TPTGGTVCTT AQQNPNPGAA SQQNLDDMTL KDLMSSVEKD MRQLQAETND LVTNVYDARE YTRRAIDQIL QLVKGFERFQ K |
| **A14** | MPXV | ProteoGenix | E. coli | Asn24-Ala70 | - |
| **A26** | MPXV | ProteoGenix | E. coli | Asp2-Glu75 | - |
| **A27** | MPXV | ProteoGenix | E. coli | Leu428-Thr695 | - |
| **A27** | VACV | SinoBiological | E. coli | Met1-Glu110 | - |
| **A29** | MPXV | ProteoGenix | Mammalian cells | Asp2-Glu110 | - |
| **A33** | MPXV | SinoBiological | E. coli | - | - |
| **A33** | VACV | SinoBiological | E. coli | Val57-Asn185 | - |
| **A35** | MPXV | SinoBiological | Mammalian cells (HEK293 Cells) | - | - |
| **A36** | MPXV | ProteoGenix | E. coli | Ile29-Lys168 | - |
| **A44** | MPXV | ProetoGenix | E. coli | Asp2-Thr74 | - |
| **B2** | MPXV | Abbexa | Mammalian cells | Ser19-Asp274 | SP QTSKKIGDDA TISCSRNNTN YYVVMSAWYK EPNSIILLAA KSDVLYFDNY TKDKISYDSP YDDLVTTITI KSLTAGDAGT YICAFFMTST TNDTDKVDYE EYSIELIVNT DSESTIDIIL SGSTPETISE KPEDIDNSNC SSVFEIATPE PITDNVEDHT DTVTYTSDSI NTVNASSGES TTDETPEPIT DKEEDHTVTD TVSYTTVSTS SGIVTTKSTT DDADLYDTYN DNDTVPPTTV GGSTTSISNY KTKD |
| **B5** | MPXV | ProteoGenix | E. coli | Asn26-Thr371 | - |
| **B5** | VACV | SinoBiological | Mammalian cells (HEK293 Cells) | Tyr18-His279 | - |
| **B6** | MPXV | ProteoGenix | Mammalian cells | Thr20-His279 | - |
| **C15** | MPXV | ProteoGenix | Mammalian cells | Met1-Pro176 | - |
| **C18** | MPXV | ProteoGenix | E. coli | Met1-Leu635 | - |
| **C19** | MPXV | Abbexa | E. coli | Met1-Ile372 | MWPFASVPAG AKCRLVETLP ENMDFRSDHL TTFECFNEII TLAKKYIYIA SFCCNPLSTT RGALIFDKLK EVSEKGIKII |

| | | | | | |
|---|---|---|---|---|---|
| | | | | | VLLDERGKRN LGELQSHSPD INFITVNIDK KNNVGLLLGC FWVSDDERCY VGNASFTGGS IHTIKTLGVY SDYPPLATDL RRRFDTFKAF NSAKNSWLNL CSAACCLPVS TAYHIKNPIG GVFFTDSPEH LLGYSRDLDT DVVIDKLKSA KTSIDIEHLA IVPTTRVDGN SYYWPDIYNS IIEAAINRGV KIRLLVGNWD KNDVYSMATA RSLDALCVQN DLSVKVFTIQ NNTKLLIVDD EYVHITSANF DGTHYQNHGF VSFNSIDKQL VSEAKKIFER DWVSSHSKSL KI |
| **D13** | MPXV | ProteoGenix | E. coli | Met1-Glu315 | - |
| **D14** | MPXV | ProteoGenix | Mammalian cells | Tyr20-Ala216 | - |
| **E8** | MPXV | ProteoGenix | Mammalian cells | Pro2-Lys274 | - |
| **F3** | MPXV | ProteoGenix | E. coli | Met1-Phe153 | - |
| **H3** | MPXV | SinoBiological | Mammalian cells (HEK293 Cells) | - | - |
| **I1** | MPXV | SinoBiological | E. coli | - | - |
| **L1** | MPXV | ProteoGenix | E. coli | Met1-Gln152 | - |
| **L4** | MPXV | Native Antigen Company | E. coli | - | MSLLLENLIE EDTIFFAGSI SEYDDLQMVI AGAKSKFPRS MLSIFNIVPR TMSKYELELI HNENITGAMF TTMYNIRNNL GLGDDKLTIE AIENYFLDPN NEVMPLIINN TDMTTVIPKK SGRRKNKNMV IFRQGSSPIL CIFETRKKIN IYKENMESVS TEYTPIGDNK ALISKYAGIN ILNVYSPSTS MRLNAIYGFT NKNKLEKLST NKELESYSSS PLQEPIRLND FLGLLECVKK NIPLTDIPTK D |
| **M1** | MPXV | SinoBiological | Mammalian cells (HEK293 Cells) | - | - |
